# Direct detection of humoral marker corelates of COVID-19, glycated HSA and hyperglycosylated IgG3, by MALDI-ToF mass spectrometry

**DOI:** 10.1101/2021.07.08.21260186

**Authors:** Ray K Iles, Jason K IIes, Raminta Zmuidinaite, Anna Gardiner, Jonathan Lacey, Stephen Harding, Jonathan Heeney, Helen Baxendale

**Affiliations:** MAPSciences, The iLab, Stannard Way, Bedford, MK44 3RZ, UK; Laboratory of Viral Zoonotics, Department of Veterinary Medicine, University of Cambridge, Madingley Road, CB3 0ES, Cambridge, UK; NISAD, Sundstorget 2, 252-21 HELSINGBORG, Sweden; The Binding Site Group Ltd, 8 Calthorpe Road, Edgbaston, Birmingham B15 1QT, UK; DIOSynVax, University of Cambridge, Madingley Road, CB3 0ES, Cambridge, UK; Royal Papworth Hospital NHS Foundation Trust, Cambridge, UK

**Keywords:** COVID-19, MALDI-ToF MS, plasma, glycated Albumin, Glycovariant/glycated IgG3

## Abstract

The prefusion Spike protein of SARS-CoV2 binds advanced glycation end product (AGE) glycated human serum albumin (HSA) and a higher mass, hyperglycosylated/glycated, IgG3, as determined by matrix assisted laser desorption mass spectrometry (MALDI-ToF MS). We set out to investigate if the total blood plasma of patients who had recovered from acute respiratory distress as a result of COVID-19, contained more glycated HSA and higher mass (glycosylated/glycated) IgG3 than those with only clinically mild or asymptomatic infections. A direct dilution and disulphide bond reduction method was development and applied to plasma samples from SARS-CoV2 seronegative (N = 30) and seropositive (N = 31) healthcare workers and 38 convalescent plasma samples from patients who had been admitted with acute respiratory distress syndrome (ARDS) associated with COVID-19.

Patients recovering from COVID-19 ARDS had significantly higher mass, AGE-glycated HSA and higher mass IgG3 levels. This would indicate that increased levels and/or ratios of hyper-glycosylation (probably terminal sialic acid) IgG3 and AGE glycated HSA may be predisposition markers for development of ARDS as a result of COVID-19 infection. Furthermore, rapid direct analysis of plasma samples by MALDI-ToF MS for such humoral immune correlates of COVID-19 presents a feasible screening technology for the most at risk; regardless of age or known health conditions.

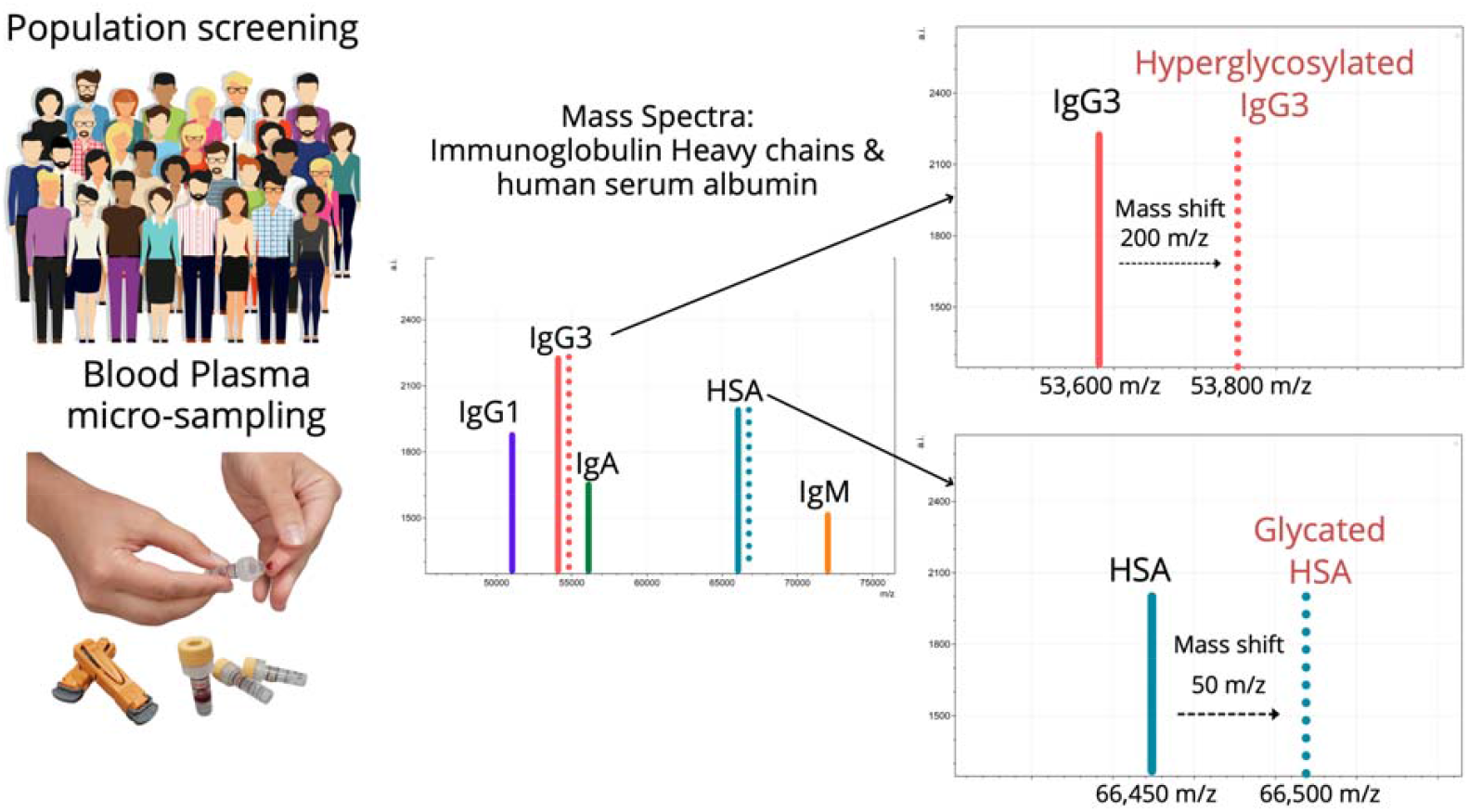

## Introduction

As the COVID-19 pandemic has gripped the world, the medical technology strategy to deal with the dynamic situation has evolved to include mass screening and mass vaccination, additional to extensive investigations to identify individuals at risk of severe disease. Mass screening has proved challenging given the logistics involved and need for affordable, easy, repeat testing, track and trace [1]. Vaccination has made a huge impact on infection transmission and hospitalisations in countries where over 70% of the adult population has been immunised [2]. Booster vaccinations are likely needed to deal with emerging variant forms of SARS CoV-2 is a new long-term realisation [3]. A huge amount of money, time and national resources have been invested in medical technologies to meet the ongoing demand for diagnostics. Given that the infection with SARS-CoV2 virus was life threatening to minority of the population [4,5] and that vaccination may reduce this risk further; there are growing calls to open back up social interaction, business, trade and international travel [6]. How this can be justified remains an ethical and global-social/political dilemma. In response to this changing focus, clinicians and scientist are seeking to identify those who are at the greatest risk of severe disease to help inform and rationalise targeted testing and booster vaccinations [7].

A range of medical technologies are being explored and developed to help inform this targeted or precision medicine approach. Currently, the focus has been on artificial intelligence (AI) identifying the “at risk” from medical notes and pre-existing test results via algorithms trained through machine learning [8,9,10]. However, whether this will be sufficient to give the level of, sensitivity and specificity, “precision” required in identifying all that are at risk is questionable. The fault will lay not with the machine learning (ML) or AI, but that insufficient robust correlating test results are being provided to the algorithms, where the diagnostic accuracy of each of the correlate will be amplified by ML. New significant and non-subjective evaluations will be required such as biomarkers and immune correlates [11].

We have recently shown that the prefusion spike protein of SARS-Cov2 binds glycated albumin and a higher molecular mass, due to glycation or hyper glycosylation, IgG3. This we proposed benefits the virus immune evasion strategies and may contribute to COVID-19 pathologies as a result of a selective cloud of serum proteins triggering thrombolytic and detrimental inflammatory pathways [12,13]. Furthermore, an increased ratio of IgG3 compared to IgG1 in the humoral response to SARS-Cov-2 is associated with an adverse outcome of a COVID-19 infection [14]. The possibility is that elevated glycated albumin along with elevated levels of IgG3 characterised by an increased molecular mass (due to glycation/glycosylation) may act as biomarkers of susceptibility to develop ARDS as a result of SARS-CoV2 infection.

We examined the mass spectra of samples from convalescent patients and sero-negative heath care workers (HCW), without immunocapture, to see if direct MALDI-ToF of diluted and disulphide reduced blood plasma would reveal gross characteristic differences in resolved immunoglobulin heavy chains and (glycated) albumin spectra. Given the logistic of simple pin prick blood home sampling, high throughput low cost MALDI-ToF mass spectrometry could provide one of the marker tests for the “at risk” population.

## Material and Methods

### Samples

Serum and plasma samples were obtained from Health care workers (HCWs) and patients referred to the Royal Papworth Hospital for critical care. COVID-19 patients hospitalised during the first wave and as well as NHS healthcare workers working at the Royal Papworth Hospital in Cambridge, UK served as the exposed HCW cohort (Study approved by Research Ethics Committee Wales, IRAS: 96194 12/WA/0148. Amendment 5). NHS HCW participants from the Royal Papworth Hospital were recruited through staff email over the course of 2 months (20^th^ April 2020-10^th^ June 2020) as part of a prospective study to establish seroprevalence and immune correlates of protective immunity to SARS-CoV-2. Patients were recruited in convalescence either pre-discharge or at the first post-discharge clinical review. All participants provided written, informed consent prior to enrolment in the study. Sera from NHS HCW and patients were collected between July and September 2020, approximately 3 months after they were enrolled in the study.

For cross-sectional comparison, representative convalescent serum and plasma samples from seronegative HCWs, seropositive HCW and convalescent PCR-positive COVID-19 patients were obtained. The serological screening to classify convalescent HCW as positive or negative was done according to the results provided by a CE-validated Luminex assay detecting N-, RBD- and S-specific IgG, a lateral flow diagnostic test (IgG/IgM) and an Electro-chemiluminescence assay (ECLIA) detecting N- and S-specific IgG. Any sample that produced a positive result by any of these assays was classified as positive. Thus, the panel of convalescent plasma samples (3 months post-infection) were grouped in three categories: A) Seronegative Staff (N = 30 samples) B) Seropositive Staff (N = 31 samples); C) Patients (N = 38 samples) [15].

### Sample Analysis by MALDI-ToF Mass spectrometry

Mass spectra were generated using a 15mg/ml concentration of sinapinic acid (SA) matrix. One µl of plasma was diluted in 40µl of ultra-pure mass spec grade water (Romil Ltd, Water beach Cambridgeshire UK). After thorough vortex mixing, 10µl of the diluted plasma was mixed with mixed 1:1 with 10µl of 20mM tris(2-carboxyethyl)phosphine (TCEP) (Sigma-Aldrich, UK) in ultra-pure water. After incubation for 15mins at room temperature 1µl of the diluted samples were taken and plated on a 96 well stainless-steel target plate using a sandwich technique [16]. The MALDI-ToF mass spectrometer (microflex® LT/SH, Bruker, Coventry, UK) was calibrated using a 2-point calibration of 2mg/ml bovine serum albumin (2+ ion 33,200 m/z and 1+ion 66,400 m/z) (Pierce™, ThermoFisher Scientific). Mass spectral data were generated in a positive linear mode. The laser power was set at 65% and the spectra was generated at a mass range between 10,000 to 200,000 m/z; pulsed extraction set to 1400ns.

A square raster pattern consisting of 15 shots and 500 positions per sample was used to give 7500 total profiles per sample. An average of these profiles was generated for each sample, giving a reliable and accurate representation of the sample across the well. The raw, averaged spectral data was then exported in a text file format to undergo further mathematical analysis.

### Spectral Data processing

Mass spectral data generated by the MALDI-ToF instrument were uploaded to an open-source mass spectrometry analysis software mMass™ [17], where it was processed by using; a single cycle, Gaussian smoothing method with a window size of 300 m/z, and baseline correction with applicable precision and relative offset depending on the baseline of each individual spectra. In the software, an automated peak-picking was applied to produce peak list which was then tabulated and used in subsequent statistical analysis.

### Statistical analysis

Peak mass and peak intensities were tabulated in excel and plotted in graphic comparisons of distributions for each antigen capture and patient sample group. Means and Medians were calculated and, given the asymmetric distributions found, non-parametric statistics were applied, such as Mann Whitney U test, when comparing differences in group distributions.

## Results

The developed direct plasma sample MALDI-ToF mass spectra analysis method was rapid and required minimal reagents, optimised pulse extraction allowed for the IgG subtypes to be resolved in most samples. From previous spectra calibration using pure preparation IgG1 heavy chains averaged at peak apex ∼51,000m/z, IgG3 at ∼54,000m/z, IgA at ∼56,000m/z and IgM at ∼74,000m/z. Human Albumin was at 66,400m/z (1+) and Transferrin at 79,600m/z. All are broad heterogenous peaks reflecting glycosylation/glycation & sequence variation, and were identifiable in the plasma sample spectra (see figure 1). A further as yet to be confirmed Ig heavy chain (IgX), presumed to be IgG4, was often seen resolving at ∼48,000m/z [12,13].

**Figure 1 -.**
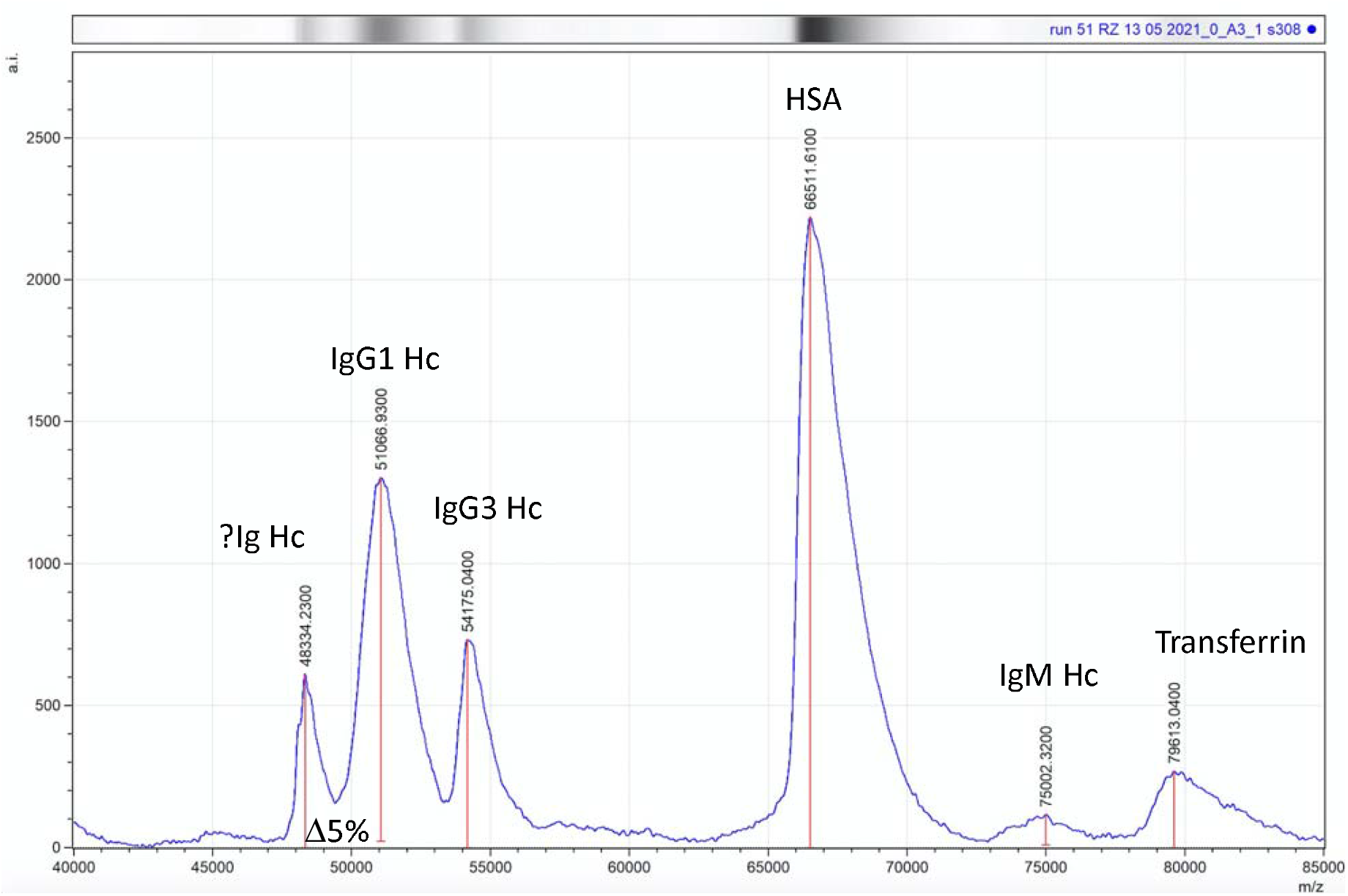
Mass spectra profile, 40,000m/z to 85,000m/z, of human reduced plasma samples (treated with TCEP) in order to reveal Immunoglobulin heavy chains (IgM Hc ‘75,000m/z, IgA ∼ 56,000 m/z not found, IgG3 54,000 and unconfirmed IgG(x), thought to be IgG4, at 48,000 m/z. Human serum albumin resolved at 66,400+ and transferrin at 79,000m/z.

### Direct plasma total IgG1 spectra

IgG1Hc was detected in all samples and was the most dominant and abundant immunoglobulin such that less abundant IgG3 and the related Ig heavy chain termed IgX Hc were often masked under the broad polyclonal IgG1 peak. Although intensity levels varied no significant association with the COVID-19 sample sets was found. Similarly, variance in IgG1 average molecular mass, although slightly lower for the ARDS COVID-19 patient set, was not statistically significant: Mean difference Δ31m/z, -0.06% change in mass, not significant *p=0*.*177*. (See figure 2)

**Figure 2 –.**
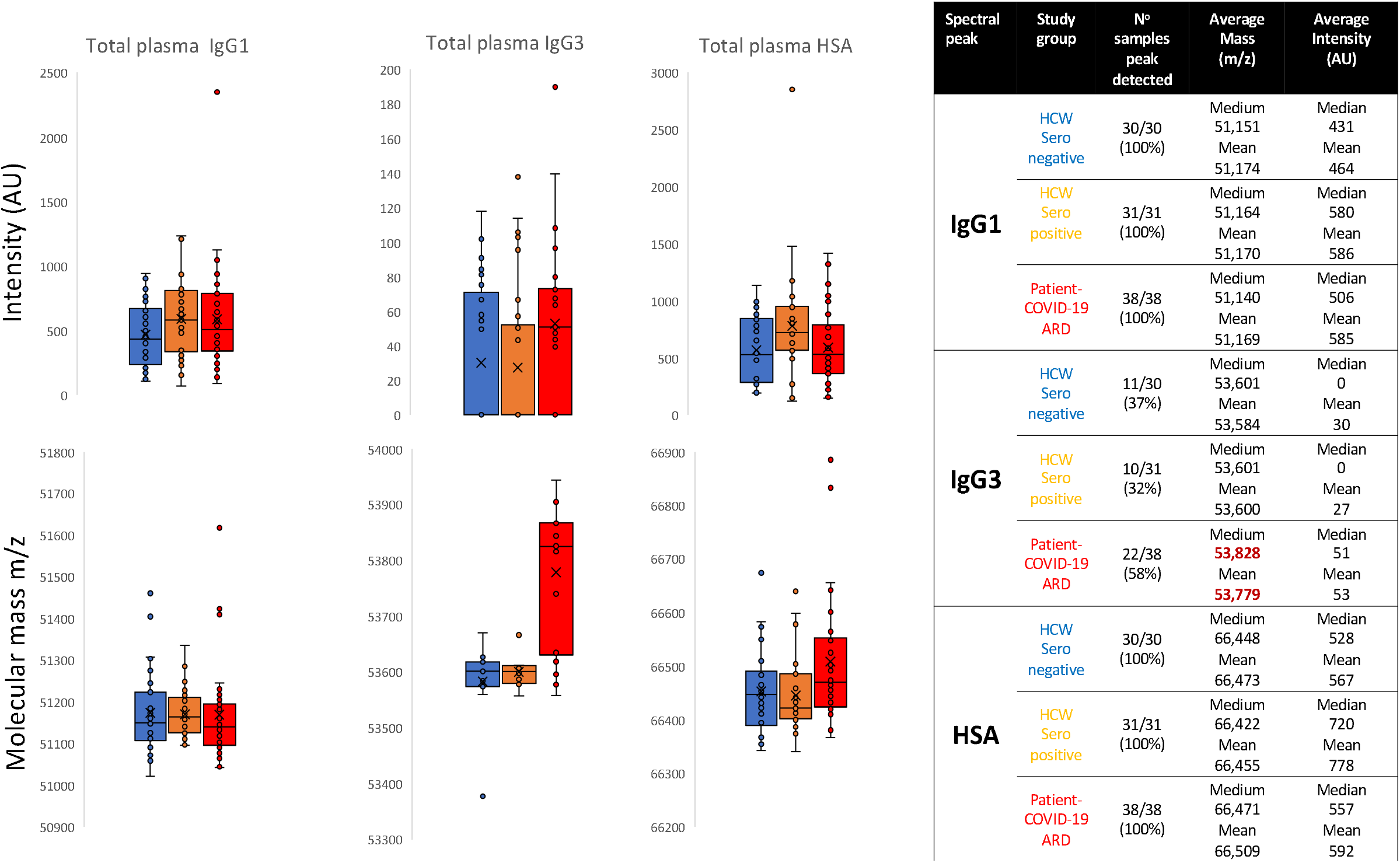
Box and whisker plots of relative intensities and variance in peak apex molecular mass of IgG1 heavy chains (IgG1 Hc), IgG3 heavy Chains (IgG3 Hc) and human Serum albumin (HAS) for the different sample groups: **Blue** represents data from SARS-CoV2 sero-negative HCW, **Orange** from SARS-CoV2 HCW sero-positive having recovered from COVID-19 with mild symptoms and **Red** sample data from convalescent patients recovering from COVID-19 ARDS. The table to the right is numeric data from the plots detailing the mean and medium values of peak intensity (AU) and molecular mass (m/z) of IgG1Hc, IgG3Hc and HSA for the respective groups.

### Direct plasma total IgG3 spectra

IgG3 Hc mass spectral peaks were detected in 43/99 samples and showed an increasing intensity in those who had been infected with SARS-CoV2, with highest levels being found in the ARDS-COVID-19 patients’ convalescent plasma: In the sero-negative HCW samples, peak detected in 37% of samples, mean intensity 30AU; sero-positive HCW samples, peak detected in 32% of samples, mean intensity 27AU and COVID-19 ARDS patient samples peak detected in 58% of samples, mean intensity 53AU. The molecular mass of the total IgG3 Hc in the plasma samples of the ARDS-COVID-19 patient convalescent plasma was significantly higher than the other groups (Mean difference: Δ200m/z, +0.37% change in mass, *p=*0.00236) (see figure 2)

### Direct plasma HSA spectra

Human Albumin was detected in all samples and no statistically significant variance in albumin Intensity could be found in these post infection convalescence plasma samples. However, the molecular mass of the convalescent plasma HSA in the patients who were recovering from ARDS as a result of SARS-CoV2 was statistically significantly larger than total HSA found in HCWs who had recovered from SARS-COV2 infection with only mild symptoms: Mean difference Δ50m/z, +0.08% change in mass, *p*=0.00758).

## Discussion

That the total HSA in patients who had developed ARDS COVID-19 had an average molecular weight higher than those developing only mild symptoms, is a significant clinical biomarker finding. But not entirely surprising, given that the elderly and particularly those with diabetes are most at risk [18]; and our previous study that the prefusion complete Spike protein binds AGE/glycated albumin [12]. Thus, it is entirely consistent with the finding of higher molecular weight glycated albumins are at such a high proportion in unextracted plasma that it skews the average mass of HSA measured, in those who had developed ARDS as a result of COVID-19.

More surprising was that, when detected, the MALDI-ToF mass spectra of total IgG3Hc was also significantly higher in the COVID-19 ARDS convalescent plasma (see figure 2). It had been reported that a dominance of anti-SARS-CoV2 IgG3 occurs in the immune responses to SARS-CoV2 in those who died from COVID-19 [14]. We had shown an increase in the mass of IgG3Hc bound to SARS-CoV2 prefusion Spike protein and eluted from magnetic bead to which it was conjugated [13]. However, to find a dominance of this higher molecular weight IgG3Hc in total plasma from patients who had recovered from COVID-19 ARDS indicates this is probably a characteristic of their inherent immunoglobulin synthesis, i.e., hyper glycosylation [19, 20]. Further to being a measurable risk biomarker, the magnitude of the mass difference at ∼200m/z is larger than that previously encountered when extracting SARS-CoV2 antigen bound antibodies (medium increase ∼150m/z), and better resolved [13].

This increased resolution of a higher mass/ hyper-glycosylated total IgG3Hc, compared to when specific IgG3 capture and elution was analysed by MALDI-ToF mass spectra, could be explained by terminal glycan sugar hydrolysis and loss from the N- and O-linked moieties. Sialic acid is a common terminal structure on all glycans; however, several factors result in loss of detectable sialic acid residues including low pH and/or high temperature [21 – 25]. This is particularly common in MALDI-ToF mass spectrometry with acidic matrices (such as sinipinic acid - SA) [26,27]; and certainly, will have occurred to a large extent during our extraction of anti SARS-CoV2 Immunoglobulins from antigen bound magnetic beads with 5% acetic acid [12,13]. Indeed, we have noted that when recovering bound albumin 5% acetic acid was sufficient to hydrolyse the N-terminal aspartic acid residue from the albumin, resulting in a mass decrease of ∼115m/z compared to HSA not exposed to 5% acetic acid. Such prolonged acetic acid hydrolysis at elevated temperatures will selectively cleave at all aspartic acid residues within a protein [28].

Terminal sialyation of glycan branches have been implicated in many regulatory processes of glycoprotein including circulatory half-life [29]. Terminal sialyation of O-linked glycan of the carboxy terminal region of the beta-subunit of human chorionic gonadotropin are responsible for this hormone’s increased blood circulatory half-life to 24hrs compared to its 82% homologous hormone Luteinising hormone which has a half-life of only ∼20mins [30-32]. IgG3 has a reported variable half-life and is the only IgG subtype to have serine O-linked glycosylation sites within the defining neck region of the common structure 9 [33] (see figure 3). The variability and extent to which these serine residues of the IgG3 neck domain are glycosylated and terminally sialyation may be decided by other factors including a genetic preference. Similarly, IgG3 has an additional potential N-linked glycosylation site within the neck domain which may or may not be so processed [33]. Furthermore, tri-antennary rather than bi-antennary branched glycans are a common feature of variant glycosylation [34,35]. Thus, the loss of terminal sialic acids and other chain saccharides by acidic hydrolysis may mean that only residual branching sugars in extracted and purified immunoglobulin peptide chains are left to, poorly, reflect the true extent of glycan variances [24]. The direct analysis of unextracted plasma by MALDI-ToF circumvents sialic acid and other glycan losses, due to acid hydrolysis, which are an unintended consequence of purification. Thus, explaining the much clearer size differences in IgG3Hc, revealed in direct plasma sample analysis by MALDI-ToF mass spectrometry.

**Figure 3 –.**
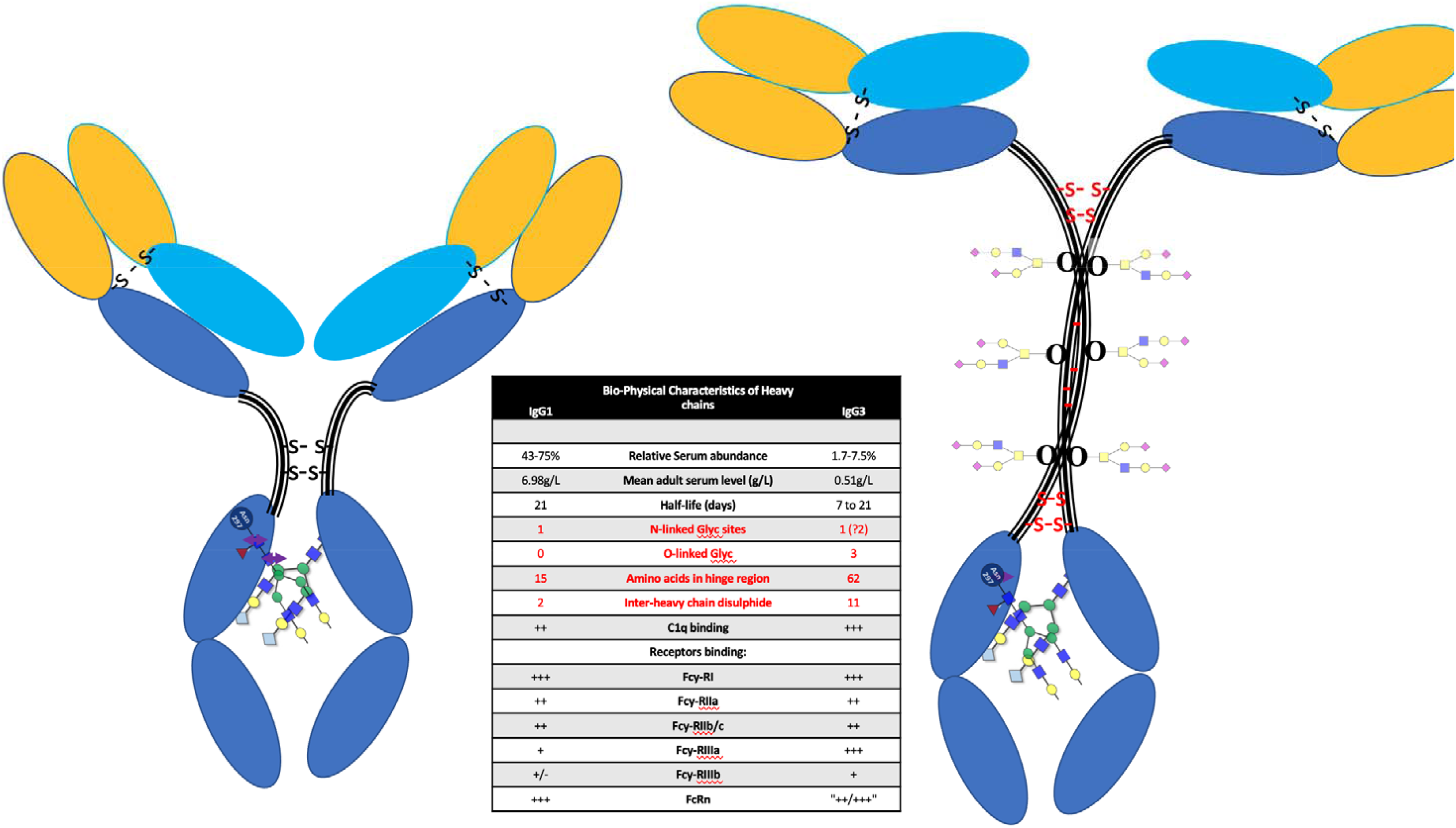
Diagrammatic representation of IgG1 and IgG3 illustrating the differences in heavy chain structure with special reference to the larger neck domain and the O-linked glycosylation sites found there.

The importance of sialic acid residues as a target for attachment of Influenza is well known [36,37]. However, human coronaviruses OC43 and HKU1 also bind to 9-*O*-acetylated sialic acids via a conserved receptor-binding site in spike protein domain A [38]. Equivalent to the S1 subunit of SARS-CoV2 this sialic acid residue binding domain may still be partially present in this corona virus. Indeed, sialic acid binding has been postulated to be the key feature of corona virus zoonotic transfer [39]. Furthermore, so strong and specific is the spike complexes binding to sialic acid that one group has used terminally sialyated glycans as a solid phase for SARS-CoV2 capture in a lateral flow diagnostic device [40]. It is not a large binding domain and may be a charge specific attraction. Terminal sialic acids confer a strong net negative charge, whilst other terminal glycan saccharides do not. Similarly, some of the AGE glycation product deposit a structured negative charge to albumin [41]. This may be a major component of the biochemistry of glycated HSA and hyperglycosylated (and/or glycated) IgG3 binding by SARS-CoV2 prefusion Spike protein.

The human cost of the COVID-19 pandemic has been nearly four million lives lost worldwide. The financial cost to the USA is estimated at more than $16 trillion, or approximately 90% of the annual gross domestic product (GDP) of the US [42]. In the UK, borrowing (deficit) in 2021 grew by £300 billion, directly attributable to COVID-19 and in 2020 GDP decreased by 11.3% [43]. With estimates of gross domestic product lagging behind pre-pandemic forecast by 3% annually for many years, the final cost for the UK maybe a similar 90% of GDP. If this is reflected globally then 90% of the global GDP will put the final cost at $76 Trillion.

Clearly the investment in medical technologies to detect the virus, prevent its infection and identify those at most risk from COVID-19, along with future viral outbreaks, is both ethically and financially justified. In this respect clinical mass spectrometry has only just started to indicate its utility [44, 45].

The potential molecular mechanisms by which sialic acid glycosylation and alike glycations contribute to the pathogenesis of COVID-19 require further investigation. Nevertheless, the direct detection of AGE glycated HSA and high mass (hyperglycosylated and or glycated) IgG3Hc, as a rapid biomarker screening test for the identification of those most at risk of developing life-threatening ARDS as a result of SAR-CoV2 infection, is a compelling possibility.

## Data Availability

Upon request to corresponding author

## Abbreviations

HCW: Health care workers
ARDS: Acute respiratory distress syndrome
ITU: Intensive Therapy unit
AGE: advanced glycation end products
MALDI-ToF: Matrix assisted laser desorption ionization – time of flight
HSA: Human serum albumin
IgG3: Immunoglobin G subtype 3

## Acknowledgements

This study was undertaken by the Humoral Immune Correlates to COVID-19 (HICC) consortium, funded by the UKRI and NIHR; grant number G107217 (COV0170 - HICC: Humoral Immune Correlates for COVID-19). RKI is also funded by NISAD Ideell Förening (charitable association) Organisationsnummer 802528-6157

We grateful thank Bruker UK Ltd, Coventry for the loan of Microflex MALDI-Tof mass spectrometer and Dr Erika Tranfield and Dr Julie Green for technical support with Bruker MALDI-ToF.

